# AI-based prognosis and chemotherapy benefit in node-positive, HR+ breast cancer: a blinded external validation on SWOG-8814 trial

**DOI:** 10.64898/2026.10.21.26366092

**Authors:** Alastair M. Thompson, Allison Meisner, Kathy S. Albain, Cerise Tang, Dhruva Biswas, Jungkyu Park, Ken Zeng, Joseph Cappadona, Chuwen Liu, Alec McClean, Jeroen Berrevoets, Linus Bao, Bartosz Machura, Jan Witowski, Krzysztof J. Geras, Corey W. Speers, Daniel F. Hayes, Kevin Kalinsky, Priyanka Sharma, James M. Rae, Andrew K. Godwin

## Abstract

**Background:** The benefit from adjuvant chemotherapy varies among patients with postmenopausal, node-positive, hormone receptor-positive breast cancer. Genomic assays identify patients likely to benefit from chemotherapy but are expensive and tissue consuming. AI-enabled biomarkers may provide scalable alternatives. We performed a blinded external evaluation of Ataraxis Breast CTX, a causal multi-modal AI model that integrates H&E images with clinical variables, in the phase III SWOG-8814 trial.

**Methods:** The analytic cohort comprised 677 patients (46% of the 1477 enrolled) with digitized H&E slides. Two CTX scores were evaluated: a histology-only score and a multi-modal score integrating pathology features with clinical variables. CTX weights and thresholds were locked before evaluation and SWOG-8814 data were not used for retraining. Predictions were generated blinded to outcomes and provided to SWOG for validation. The primary endpoint was disease-free survival (DFS), matching the primary endpoint of the trial.

**Findings:** The histology-only score was prognostic for DFS (HR = 1.36, 95% CI = 1.22-1.51) and added prognostic information beyond clinical variables alone (p < 0.001). The multi-modal score showed a stronger association with DFS (HR = 1.60, 95% CI = 1.44-1.78, p < 0.001) and greater discrimination than the histology-only score (C-index 0.66 vs 0.59). In exploratory analyses, modeled as a continuous score, CTX-benefit showed a treatment-by-biomarker interaction of HR of 0.82 (0.64-1.04, p = 0.053) for CET versus ET. The chemotherapy effect on DFS was numerically larger in the high-benefit subgroup (n = 480, HR = 0.79, 95% CI = 0.59-1.05) compared to the low-benefit subgroup (n = 197, HR = 0.91, 95% CI = 0.49-1.68).

**Interpretation:** In this blinded validation, CTX was significantly associated with DFS, supporting its prognostic value. In exploratory analyses, CTX demonstrated suggestive evidence of a treatment-by-biomarker interaction for chemotherapy benefit. These findings support further prospective validation of CTX as a tissue-sparing tool for recurrence-risk assessment and treatment selection.

**Funding:** National Cancer Institute, The Hope Foundation for Cancer Research, and Ataraxis AI.

**Research in context:** *Evidence before this study:* We searched PubMed for studies published between Jan 1, 2016, and Sept 1, 2026, using the terms (“digital pathology” OR “deep learning” OR “artificial intelligence”) AND (“hematoxylin” OR “haematoxylin” OR “H&E”) AND “breast cancer” AND (“chemotherapy benefit” OR “predictive biomarker”), without language restrictions. Multiple AI models based on H&E-stained whole-slide images, with or without clinical variables, have been developed to estimate recurrence risk in early hormone receptor-positive (HR+) breast cancer. However, robust evidence for AI-based prediction of chemotherapy benefit remains limited as most models either infer the 21-gene Recurrence Score or estimate baseline recurrence risk. To date, no AI-based markers have been shown to be predictive of chemotherapy benefit in a fully blinded, prespecified analysis of randomized clinical trial data.

*Added value of this study:* This is a blinded, pre-specified external validation of an AI-based multi-modal biomarker in SWOG-8814, a phase III randomized chemotherapy trial in postmenopausal, node-positive, hormone receptor positive breast cancer. The AI model weights and thresholds were locked prior to evaluation, no data from the trial was used for model development, and predictions were analyzed independently by the SWOG statistical center. CTX was significantly prognostic for SWOG-8814’s primary endpoint (disease-free survival) across the full cohort, within both treatment arms, and in the 1-3 positive-node subgroup. CTX also added prognostic information beyond established clinical covariates. In exploratory analyses of chemotherapy benefit, the treatment-by-biomarker interaction did not reach statistical significance, though the magnitude of chemotherapy benefit appeared to vary by subgroup.

*Implications of all the available evidence:* These findings support CTX as an AI-based approach to recurrence-risk assessment in postmenopausal women with node-positive HR+ breast cancer, a group in whom decisions about adjuvant chemotherapy remain challenging. As CTX derives prognostic information from routinely collected H&E-stained slides, it avoids the tissue consumption, cost, and turnaround delays that limit access to genomic assays. Prospective validation in contemporary node-positive cohorts receiving taxane-based chemotherapy and aromatase inhibitors will be required before CTX-guided treatment selection can be recommended in routine practice.

## Introduction

Breast cancer is the most common cancer in women worldwide (Bray et al., 2024) and hormone receptor-positive (HR+) disease accounts for approximately 80% of cases (Early Breast Cancer Trialists’ Collaborative Group (EBCTCG), 2005; Howlader et al., 2014). Trials including SWOG-8814, have established that adding adjuvant chemotherapy to endocrine therapy improves survival outcomes. However, risk of recurrence and chemotherapy benefit varies substantially between individuals (Albain et al., 2009; Fisher et al., 1997; Kalinsky et al., 2021; Sparano et al., 2018) Genomic assays, such as the 21-gene Recurrence Score (RS) assay, were designed to be prognostic tools and have since been shown in large prospective randomized trials to identify patients who can safely forgo adjuvant chemotherapy (Albain et al., 2010; Kalinsky et al., 2021; Sparano et al., 2018). However, commercially available genomic assays remain expensive and tissue-consumptive, limiting their accessibility (Brufsky et al., 2025; Filipits et al., 2011; Paik et al., 2004). Digital pathology, which extracts prognostic information directly from routinely collected hematoxylin and eosin (H&E)-stained slides using artificial intelligence (AI), offers a faster, lower-cost, tissue-sparing alternative. However, these models require rigorous validation before clinical use (Howard et al., 2023; Shamai et al., 2026; Witowski et al., 2026). In addition, accurate identification of patients expected to benefit from chemotherapy is essential to guiding treatment, as overtreatment exposes patients to severe acute and long-term toxicities, financial burden, and reductions in quality of life (Shapiro & Recht, 2001; Smith et al., 2019; Zafar & Abernethy, 2013) while undertreatment risks preventable recurrence (Shapiro & Recht, 2001; Zafar & Abernethy, 2013). Therefore, digital pathology biomarkers that accurately estimate individual-level chemotherapy benefit, rather than simply recurrence risk, could help to guide therapeutic decision-making in early-stage HR+ breast cancer.

The SWOG-8814 trial was a phase III randomized controlled trial that accrued postmenopausal women diagnosed with node-positive early HR+ breast cancer randomized to either tamoxifen alone, to cyclophosphamide, doxorubicin, fluorouracil (CAF) chemotherapy followed by tamoxifen (CAF-T), or to CAF plus concurrent tamoxifen (CAFT) (Albain et al., 2009). A subsequent analysis of the trial that added the RS assay demonstrated that RS was prognostic for disease-free survival (DFS) in this population, where patients with low RS (<18) had substantially better outcomes than patients with high RS (>30) (Albain et al., 2010). Notably, the treatment-by-biomarker interaction for RS was significant in the first 5 years (p = 0.029), with no additional prediction beyond 5 years (p = 0.58). However, the cumulative benefit remained at 10 years. Results were similar for overall survival and breast-cancer-specific survival (Albain et al., 2010).

Here, we evaluate Ataraxis Breast CTX, a causal multi-modal AI test (Biswas et al., 2026), in a blinded external validation in the SWOG-8814 trial. CTX has since identified patients at higher risk of recurrence in five independent observational cohorts (n = 1,994) and in two phase III trials, UNIRAD and TAILORx (Bachelot et al., 2026; Biswas et al., 2026; Chan et al., 2026). Evidence that CTX also identifies chemotherapy benefit comes from the observational cohorts, where the treatment-by-biomarker interaction for CTX was significant across the full population (Biswas et al., 2026). In TAILORx, the interaction was significant among the randomized patients with a recurrence score of 11-25 (Chan et al., 2026). The analysis plan was approved by SWOG and locked prior to independent execution of the analysis plan by the SWOG statistical center. In this analysis, we evaluated the prognostic performance of CTX and the ability of CTX to predict chemotherapy benefit in patients with HR+, node-positive early breast cancer.

## Methods

### Study design and population

SWOG-8814 (INT-0100; NCT00929591) was a phase III, parallel, three-arm, open-label randomized controlled trial that enrolled postmenopausal women from June 1989 to July 1995, 1,477 eligible patients were included in the intention-to-treat analysis(Albain et al., 2009). Eligible participants were postmenopausal women diagnosed with pathological stage T1-3, N1-2 (node-positive) infiltrating breast carcinoma that was estrogen-receptor and/or progesterone-receptor positive by local institutional standards. Treatment was assigned irrespective of tumor genomics. Patients were randomized in a 2:3:3 ratio to tamoxifen alone, CAF-T or CAFT, with tamoxifen given for 5 years and CAF for six cycles. In the present analysis, patients who were randomized to tamoxifen alone are denoted ET and patients randomized to CAF-T or CAFT are denoted CET. While CAF-T and CAFT were found to have differential benefit in the primary SWOG-8814 analysis (Albain et al., 2009), both chemotherapy arms were combined in this analysis as CTX was designed to predict overall chemotherapy benefit.

For the current analyses, we included 677 patients who had digitized H&E-stained slides and outcomes available (46% of the total trial population). There was no statistically significant difference in prognosis between included and excluded patients (Wald test p = 0.08) and covariates were well balanced (all standardized mean difference [SMD] < 0.10), except grade (SMD = 2.80), where the majority of excluded patients did not have grade recorded. Slides were scanned using an Aperio ScanScope scanner at 40x magnification. Of these 677 patients, 253 (37.4%) were randomized to the CAF-T arm, 257 (38.0%) were in the CAFT arm, and 167 (24.7%) were treated with tamoxifen alone. Patient inclusion, exclusion, and treatment assignment are summarized in a REMARK diagram in **Figure 1**(McShane et al., 2005). Median follow-up in the analytic cohort was 9.59 years. In this analysis, patients with unknown tumor grade (n = 77) were retained and coded as a separate “Unknown” category and patients with missing positive-node count (n = 3) were assigned the analytic cohort’s median value (3). Baseline characteristics of the cohort are described in **Table 1**.

**Figure 1.**
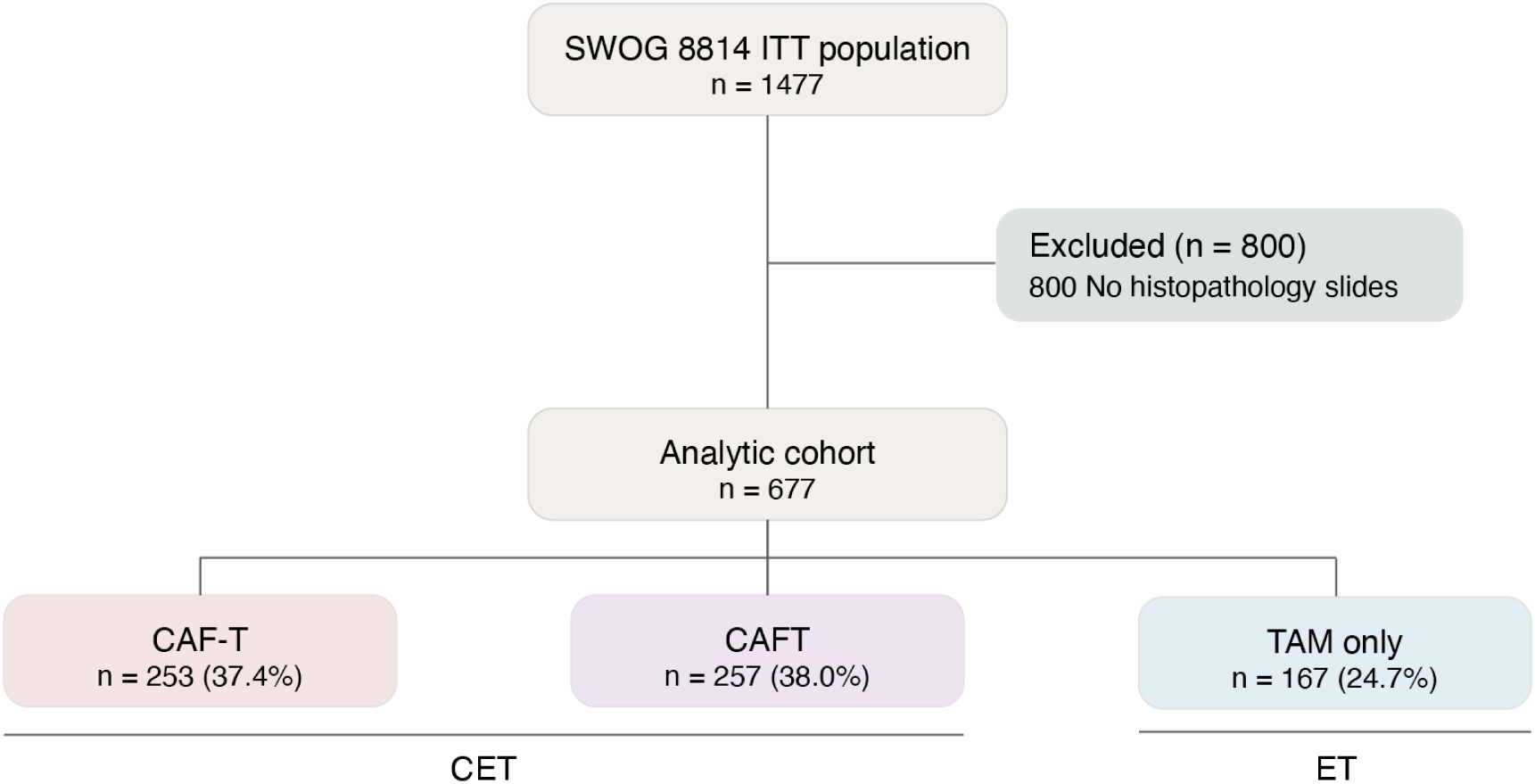
REMARK diagram. Flowchart of patients from the SWOG-8814 intention-to-treat population (N = 1477) through exclusions to the analytic cohort used for this analysis, stratified by treatment arm. REMARK denotes REporting recommendations for tumor MARKer prognostic studies; CAF-T, CAF chemotherapy followed by tamoxifen; CAFT, concurrent CAF chemotherapy and tamoxifen; TAM only, tamoxifen only; ITT, intention-to-treat.

**Table 1.**
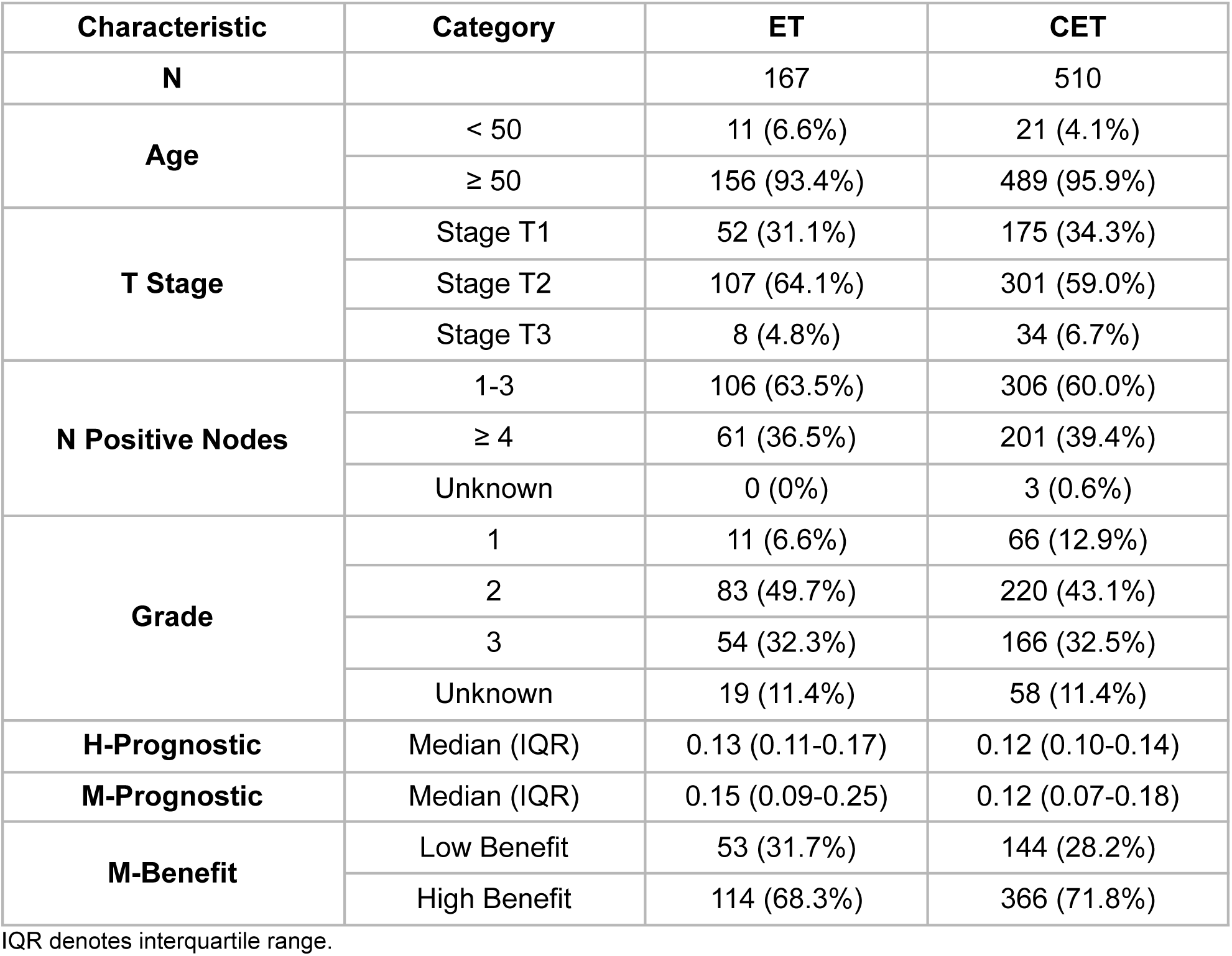
Baseline characteristics of the analytic cohort by treatment group.

### CTX development

Ataraxis Breast CTX is a causal multi-modal AI test that integrates morphologic features extracted from whole-slide images of H&E-stained pathology slides with clinical variables to estimate recurrence risk and individualized chemotherapy benefit(Biswas et al., 2026). CTX divides whole-slide images into patches and uses a pathology foundation model to encode them into feature embeddings(Cappadona et al., 2024). The slide embeddings are then combined with a clinical embedding derived from baseline clinical variables (including T-and N-stage, receptor status, and histology). The resulting joint representation is then passed to a causal model, which outputs calibrated 5-year probabilities of recurrence-free interval under counterfactual treatments: endocrine therapy alone (ET) and chemoendocrine therapy (CET). Chemotherapy benefit for each patient is defined as the difference in estimated recurrence probability when treated with chemoendocrine therapy compared to endocrine therapy alone. CTX was trained on 16,917 patients from 22 independent cohorts and was locked (including benefit subgroup thresholds) after development and not modified for this study. CTX was developed to estimate treatment effects, corresponding to the C2 intended-use category (predictive of benefit from a specific therapy) in the EBAI framework(Aldea et al., 2025).

### Definition of CTX scores

As originally developed, CTX combines information from H&E-stained slides and baseline clinical variables and also generates predictions from the slides alone (Biswas et al., 2026). In this analysis, we used both: 1) the histology (H)-score, derived from H&E-stained slides alone, and 2) the multi-modal (M)-score, which was computed using both H&E-stained slides and baseline clinical variables. The M-score corresponds to the standard CTX output and is equivalent to the CTX scores evaluated in a previous analysis of the TAILORx trial (Chan et al., 2026); we refer to it as the M-score only to distinguish it from the H-score.

CTX-prognostic, addresses how likely the patient is to recur on the treatment they received (either ET or CET) and was derived for both H-scores and M-scores, which are termed H-prognostic and M-prognostic respectively. The second, CTX-benefit, addresses how much adding chemotherapy reduces the patient’s risk of recurrence. It is the difference in predicted recurrence probability under ET and CET, where a larger score indicates greater predicted benefit. CTX-benefit was only derived from the multi-modal model (M-benefit). CTX-prognostic and CTX-benefit are related but not interchangeable as a patient may have high predicted recurrence risk yet little predicted benefit from chemotherapy.

In this study, H-prognostic, M-prognostic and M-benefit scores were modeled continuously. In addition, a pre-specified threshold of 6.5%, consistent with previous work(Bachelot et al., 2026; Tang et al., 2026) was used to dichotomize patients into M-prognostic-high and M-prognostic-low categories. Patients were also dichotomized into M-benefit-high and M-benefit-low categories using a threshold of 2% benefit in 5-year recurrence-free probability, following the cut-off used in past studies(Bachelot et al., 2026; Biswas et al., 2026; Chan et al., 2026).

### Study objectives

The endpoints and objectives of this analysis were pre-specified in a statistical analysis plan drafted by Ataraxis AI, revised by SWOG investigators, and jointly approved. The primary endpoint used in this study was disease-free survival (DFS), while overall survival (OS) was evaluated as a secondary endpoint. OS analyses were conducted by Ataraxis AI after outcome data were released by SWOG. A total of 280 DFS and 209 OS events were recorded in the analytic cohort. Disease-free survival was defined in the SWOG-8814 trial as recurrence or death from any cause. The first objective was to evaluate whether H-prognostic and M-prognostic were significantly associated with DFS using three Cox proportional hazards models (H-prognostic alone, H-prognostic with clinical covariates, and M-prognostic alone). The clinical covariates evaluated in the H-prognostic with clinical covariates model were grade, age at diagnosis, and number of positive nodes. Receipt of adjuvant chemotherapy was included in the adjusted covariates in the models which included all patients. Hazard ratios are reported per 1 standard deviation (SD) difference in score. In the model with H-prognostic and clinical covariates, we tested the one-sided hypothesis that the H-prognostic with clinical covariates model added prognostic information independent of the clinical variables, using a one-sided likelihood ratio test, based on halving the χ² p-value.

To assess whether prognostic performance varied over time, we additionally fitted Cox models stratified by follow-up interval, estimating hazard ratios for events occurring within the first 5 years and, separately, for late events occurring beyond 5 years among patients who remained event-free at 5 years. Finally, we evaluated the discriminatory ability of H-prognostic and M-prognostic for DFS using Cox proportional hazards models, Harrell’s C-index, and time-dependent area under the receiver operating characteristic curve (AUROC) estimated at 5 years. Kaplan-Meier curves were estimated for M-prognostic groups, confidence intervals were calculated using Greenwood’s formula, and p-values were calculated using two-sided Wald tests. Prognostic performance was assessed separately in patients who received ET, in patients who received CET, and in patients with 1-3 positive nodes (exploratory).

The second objective was to determine whether the effect of chemotherapy differed by predicted benefit estimated by M-benefit. This was evaluated in an exploratory analysis by fitting a Cox model with a treatment-by-biomarker interaction for M-benefit, where M-benefit was modeled as a continuous variable, standardized to mean zero and standard deviation one. This was tested using a one-sided likelihood ratio test, under the alternative hypothesis that treatment benefit increases as M-benefit increases (*i.e.,* that the interaction hazard ratio was less than one). The interaction is reported as the ratio of CET-vs-ET hazard ratios per 1 SD increase in M-benefit score; values less than 1 indicate greater relative benefit of chemotherapy at higher M-benefit. We also report the CET-vs-ET hazard ratio within M-benefit-low and M-benefit-high groups, along with 5-and 10-year estimates of DFS. For the M-benefit-low group, we report a two-sided Wald test, reflecting the expectation of no treatment difference and, for the M-benefit-high group, we used a one-sided Wald test, reflecting the expectation that patients receiving chemotherapy have better survival.

## Results

### Prognostic evaluation of pathology-only and multi-modal models

H-prognostic was associated with DFS (HR = 1.36, 95% CI = 1.22-1.51, p < 0.001) and remained so after adjustment for clinical covariates (HR = 1.26, 95% CI = 1.11-1.43, p < 0.001). M-prognostic showed a stronger association with DFS with an HR of 1.60 (95% CI = 1.44-1.78, p < 0.001; **Table 2**). Adding H-prognostic to a model containing clinical covariates alone improved model fit (likelihood ratio test p < 0.001). M-prognostic discriminated better than H-prognostic (C-index = 0.66 [95% CI = 0.62-0.69] vs 0.59 [0.56-0.63]; 5-year AUROC = 0.70 [0.66-0.75] vs 0.62 [0.57-0.67]; **Table 3**).

**Table 2.** Cox proportional hazards model performance for DFS.

| Group | Model | Hazard Ratio | 95% CI | p-value |
| --- | --- | --- | --- | --- |
| **All patients** | H alone | 1.36 | 1.22-1.51 | < 0.001 |
|  | H + clinical features | 1.26 | 1.11-1.43 | < 0.001 |
|  | M alone | 1.60 | 1.44-1.78 | < 0.001 |
| **ET only** | H alone | 1.38 | 1.18-1.62 | < 0.001 |
|  | H + clinical features | 1.41 | 1.17-1.71 | < 0.001 |
|  | M alone | 1.62 | 1.35-1.94 | < 0.001 |
| **CET** | H alone | 1.32 | 1.14-1.54 | < 0.001 |
|  | H + clinical features | 1.18 | 0.99-1.40 | 0.072 |
|  | M alone | 1.60 | 1.39-1.84 | < 0.001 |
| **1-3 Nodes** | H alone | 1.33 | 1.14-1.56 | < 0.001 |
|  | H + clinical features | 1.21 | 0.99-1.47 | 0.063 |
|  | M alone | 1.29 | 1.11-1.50 | < 0.001 |

Hazard ratios for disease-free survival per 1 SD increase in score, from Cox proportional hazards models fitted in the overall analytic cohort (n=677) and within subgroups defined by treatment received and nodal burden. H + clinical features was adjusted for grade, age at diagnosis, number of positive nodes, and receipt of adjuvant chemotherapy (the latter in the all-patients and 1-3 node models only). H denotes histology score; M, multi-modal score.

**Table 3.** Discriminatory performance for DFS.

| Subgroup | Score | Harrell's C-index | 5-Year AUROC |
| --- | --- | --- | --- |
| **All patients** | H-prognostic | 0.59 (0.56-0.63) | 0.62 (0.57-0.67) |
|  | M-prognostic | 0.66 (0.62-0.69) | 0.70 (0.66-0.75) |
| **ET only** | H-prognostic | 0.61 (0.54-0.68) | 0.64 (0.53-0.74) |
|  | M-prognostic | 0.69 (0.62-0.75) | 0.72 (0.63-0.81) |
| **CET** | H-prognostic | 0.58 (0.55-0.62) | 0.61 (0.56-0.67) |
|  | M-prognostic | 0.65 (0.61-0.69) | 0.70 (0.65-0.75) |
| **1-3 Nodes** | H-prognostic | 0.58 (0.53-0.64) | 0.62 (0.54-0.70) |
|  | M-prognostic | 0.60 (0.55-0.65) | 0.64 (0.56-0.72) |

95% confidence intervals are in brackets. H denotes histology score; M, multi-modal score; ET, endocrine therapy only; CET, chemoendocrine therapy.

Next, to determine whether M-prognostic can stratify patients by recurrence risk, we binarized patients into high-and low-risk groups. A total of 555 patients (82%) were classified as CTX high-risk while 122 (18%) were classified as low-risk. High-risk patients had a significantly lower 5-year DFS (73.4%, 95% CI = 69.8-77.1%) than low-risk patients (89.2%, 95% CI = 83.8-94.9%, **Figure 2a**). There was a statistically significant difference in DFS between the two risk groups across the follow-up period (p < 0.001). High-risk patients also had a markedly lower 10-year DFS of 52.1% (95% CI = 47.7-57.0%) compared to 71.8% (95% CI = 62.9-81.9%) in low-risk patients.

**Figure 2.**
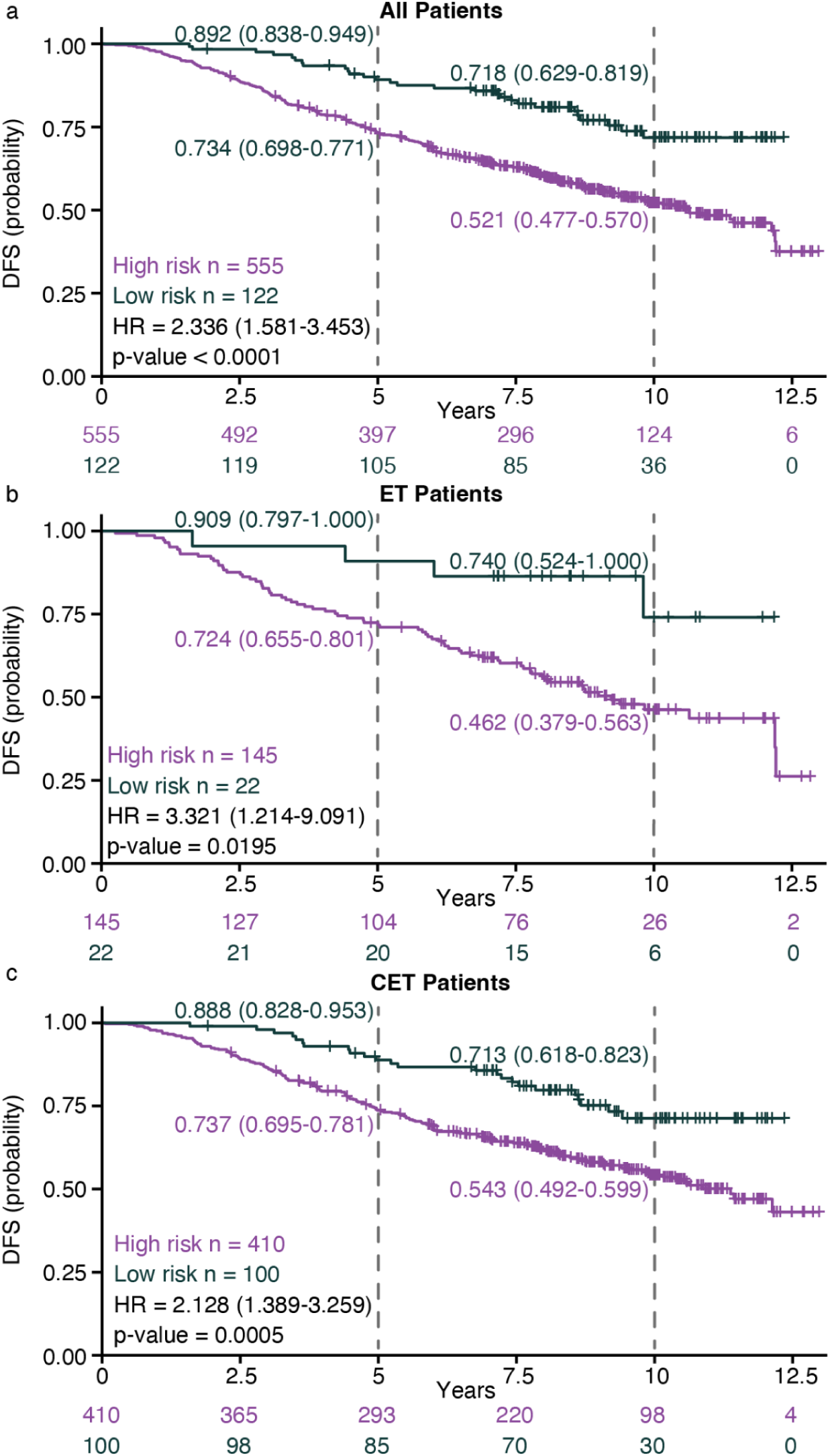
CTX demonstrated prognostic value in patients across treatment arms. Panels a, b, and c show DFS estimates in all patients (panel a), patients treated ET (panel b), and patients treated with CET (panel c). High-and low-risk groups were defined using a prespecified threshold for the M-prognostic score. Kaplan-Meier estimates of five-and ten-year DFS with 95% confidence intervals are shown for each risk group. Hazard ratios with 95% confidence intervals and p-values were derived from univariable Cox proportional hazards models (Wald test), with the low-risk group as the reference. Numbers at risk at each time point are shown below each panel. CET denotes chemoendocrine therapy; CI, confidence interval; HR, hazard ratio; DFS, disease-free survival; ET, endocrine therapy alone.

### Model performance stratified by adjuvant chemotherapy status

M-prognostic remained significantly associated with DFS in both ET (HR = 1.62, 95% CI = 1.35-1.94, p < 0.001; **Table 2**) and CET (HR = 1.60, 95% CI = 1.39-1.84, p < 0.001) arms. Discrimination was slightly stronger in patients randomized to ET (ET: M-prognostic C-index = 0.69, 95% CI = 0.62-0.75; CET: C-index = 0.65, 95% CI = 0.61-0.69; **Table 3**). Within patients treated with ET, DFS was significantly different between high-and low-risk groups (p = 0.0195; **Figure 2b**), with lower 5-year DFS observed in high-risk patients (72.4%, 95% CI = 65.5-80.1%) than low-risk patients (90.9%, 95% CI = 79.7-100%). Consistent with this, 10-year DFS was lower in high-risk patients (46.2%, 95% CI = 37.9-56.3%) than in low-risk patients (74.0%, 95% CI = 52.4-100%). DFS also differed significantly between risk groups in patients receiving CET (p = 0.0005; **Figure 2c**), where 5-year DFS was lower in high-risk patients (73.7%, 95% CI = 69.5-78.1%) than low-risk patients (88.8%, 95% CI = 82.8-95.3%). A similar pattern was observed for 10-year DFS, which was lower among high-risk patients (54.3%, 95% CI = 49.2-59.9%) than among low-risk patients (71.3%, 95% CI = 61.8-82.3%).

### Prognostic performance in patients with 1-3 positive nodes

In the 412 patients with 1-3 positive nodes, H-prognostic (HR = 1.33, 95% CI = 1.14-1.56, p < 0.001) and M-prognostic (HR = 1.29, 95% CI = 1.11-1.50, p < 0.001) were significantly associated with DFS (**Table 2**). H-prognostic demonstrated moderate discriminatory ability (C-index = 0.58, AUROC = 0.62; **Table 3**), while M-prognostic was slightly stronger (Harrell’s C-index = 0.60, AUROC = 0.64).

### Time-stratified prognostic performance

The association between H-prognostic and DFS was stronger for early events occurring in the first 5 years (n = 160 DFS events), with a hazard ratio of 1.44 (95% CI = 1.27-1.63, p < 0.001, **Table 4**) compared to a hazard ratio of 1.20 (1.00-1.45, p = 0.054) for late events (n = 120 DFS events). Similarly, the association between M-prognostic and DFS was also stronger for events within the first 5 years (HR = 1.81, 95% CI = 1.59-2.07, p < 0.001 versus HR = 1.29, 95% CI = 1.08-1.55, p = 0.006).

**Table 4.** Cox proportional hazards model performance for DFS, stratified by time.

| **Time** | **Score** | **Hazard Ratio** | **95% CI** | **p-value** |
| --- | --- | --- | --- | --- |
| **0-5 Years** | H-prognostic | 1.44 | 1.27-1.63 | < 0.001 |
|  | M-prognostic | 1.81 | 1.59-2.07 | < 0.001 |
| **> 5 Years** | H-prognostic | 1.20 | 1.00-1.45 | 0.054 |
|  | M-prognostic | 1.29 | 1.08-1.55 | 0.006 |

H denotes histology score; M, multi-modal score.

### Exploratory evaluation of CTX as a predictive biomarker for chemotherapy benefit

The exploratory treatment-by-biomarker interaction analysis, where M-benefit was modeled as a continuous variable, showed a magnitude consistent with a predictive effect but did not reach statistical significance (ratio of CET vs. ET HRs per 1 SD difference in M-benefit = 0.82, 95% CI = 0.64-1.04, p = 0.053). Using the pre-specified 0.02 threshold to define the high benefit subgroup, 480 (71%) of 677 patients were classified as M-benefit-high and 197 (29%) as M-benefit-low. In the M-benefit-low group, 5-year DFS was 90.1% (95% CI = 85.4%-95.2%) with CET and 88.7% (95% CI = 80.6%-97.6%) with ET, an absolute difference of 1.4 percentage points. At 10 years, DFS was 70.2% (95% CI = 62.1%-79.3%) in the CET arm versus 65.0% (95% CI = 49.3%-85.6%) in the ET arm. The wide confidence intervals, including for the estimated CET vs. ET HR (HR = 0.91, 95% CI = 0.49-1.68, p = 0.76; **Figure 3a**) reflects the limited sample size, and that the treatment effect was uncertain in this group. In the M-benefit-high group, 5-year DFS was 71.3% (95% CI = 66.8%-76.1%) with CET and 68.4% (60.4%-77.5%) with ET, an absolute difference of 2.9 percentage points. By 10 years the difference had widened to 10.5 percentage points (CET: DFS = 52.6%, 95% CI = 47.2%-58.6%; ET: DFS = 42.1%, 95% CI = 33.4%-53.2%), with a treatment effect that approached statistical significance (HR = 0.79, 95% CI = 0.59-1.05, p = 0.055; **Figure 3b**).

**Figure 3.**
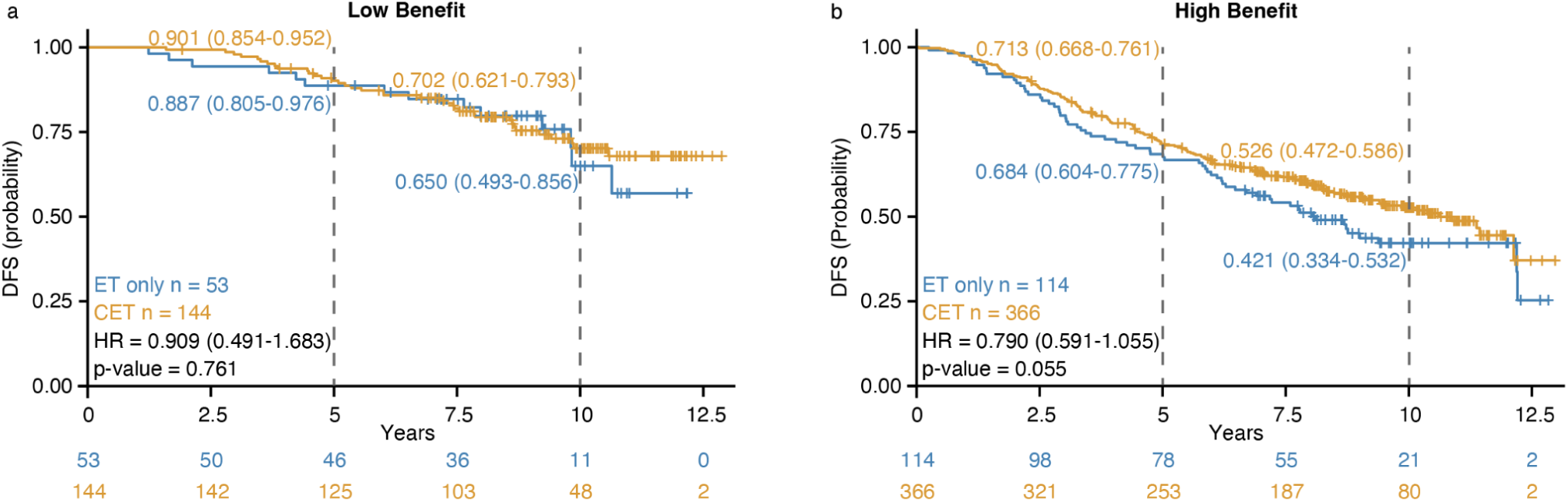
Kaplan-Meier estimates of disease-free survival by treatment group and M-benefit subgroups. DFS estimates in the M-benefit-low (panel a) and M-benefit-high (panel b) subgroups. Kaplan-Meier estimates of five-and ten-year DFS with 95% confidence intervals are shown for each risk group. Hazard ratios with 95% confidence intervals and p-values were derived from univariable Cox proportional hazards models (Wald test), with the ET only group as the reference. Numbers at risk at each time point are shown below each panel. CET denotes chemoendocrine therapy; DFS, disease-free survival; ET, endocrine therapy alone; HR, hazard ratio.

### CTX is also prognostic for overall survival

H-prognostic was associated with OS (HR = 1.40, 95% CI = 1.25-1.57, p < 0.001; **Supplemental Table 1**) and remained significant after adjustment for clinical covariates (HR = 1.32, 95% CI = 1.15-1.51, p < 0.001). M-prognostic was also significantly associated with OS (HR = 1.66, 95% CI = 1.47-1.87, p < 0.001). Discrimination was strong for both H-prognostic (Harrell’s C-index = 0.60, AUROC = 0.63; **Supplemental Table 2**) and particularly for M-prognostic (Harrell’s C-index = 0.67, AUROC = 0.73).

## Discussion

Adjuvant chemotherapy improves patient outcomes for some women diagnosed with early HR+ breast cancer, but its benefits must be weighed against acute and long-term toxicity(Smith et al., 2019). The SWOG-8814 study established that the benefit of adding chemotherapy to endocrine therapy is heterogeneous among patients with HR+, node-positive, early breast cancer. A reanalysis of SWOG-8814 found that higher recurrence scores were associated with larger chemotherapy benefit and a smaller treatment effect at low scores, although the treatment-by-biomarker interaction was not statistically significant for continuous RS(Albain et al., 2010). In addition, RS remains, at its core, an estimate of baseline risk rather than predicted benefit(Paik et al., 2004). RxPONDER (SWOG-S1007), a prospective trial designed to assess modification of chemotherapy benefit by RS in patients with node-positive disease, confirmed the RS prognostic value but did not demonstrate a treatment-by-biomarker interaction for RS in the overall study population(Kalinsky et al., 2021). Therefore, tools that directly estimate individual treatment benefit remain an unmet need.

Here, in a blinded external analysis of the SWOG-8814 trial, we evaluated whether Ataraxis Breast CTX provided prognostic information for disease-free survival and explored whether CTX-estimated CET benefit was associated with observed treatment effect. M-prognostic was significantly associated with DFS in the overall analytic cohort and within both ET and CET subgroups, with consistent discriminative performance across analyses. Prognostic performance of M-prognostic was retained among patients with 1-3 positive nodes, the most common presentation among patients enrolled in SWOG-8814 (58%). However, the effect size in this subgroup was attenuated relative to the full cohort (HR = 1.29 vs. 1.60), consistent with the lower baseline recurrence risk and fewer events in this lower-nodal-burden subgroup. M-prognostic was most strongly associated with events occurring within the first 5 years, although the association remained significant for late events, consistent with a model trained to estimate 5-year recurrence risk. In the exploratory analysis of treatment-effect modification of CET benefit by continuous M-benefit score, the continuous treatment-by-biomarker interaction for M-benefit approached but did not reach statistical significance (p = 0.053). Patients classified by M-benefit as having high-benefit (n = 480; ET = 114, CET = 366) had a numerically larger treatment effect (HR = 0.79) than those in the predicted low-benefit group (n = 197; ET = 53, CET = 144; HR = 0.91). These findings do not establish CTX as a validated predictive biomarker in this cohort but support prospective evaluation of its predictive performance in larger and more contemporary cohorts.

The prognostic association of the M-prognostic score was strongest for early events, yet the absolute DFS difference between treatment arms in the high-benefit group widened between 5 and 10 years. Chemotherapy is thought to act early and thus, the early effect may remove patients who otherwise would have recurred(Early Breast Cancer Trialists’ Collaborative Group (EBCTCG), 2005). Therefore, the curves keep separating as events continue to accrue in the endocrine-only arm. The widening at 10 years therefore likely reflects accumulated early benefit rather than late benefit of chemotherapy. However, we did not perform treatment-stratified analyses of time-varying prognostic association and numbers were small as only 21 patients in the endocrine-only, high-benefit group remained at risk at 10 years.

Several AI models have been developed to extract prognostic information from routinely collected H&E-stained slides, with many designed to infer RS or recurrence risk(Boehm et al., 2025; Howard et al., 2023; Shamai et al., 2026). However, treatment assignment can confound efforts to distinguish prognostic from predictive effects(Ballman, 2015). CTX differs from RS-inference models in its design, estimating recurrence probability under counterfactual treatment strategies rather than approximating an existing genomic score. Therefore, CTX is not constrained by the performance characteristics of the comparator assay. The prognostic performance of CTX was broadly comparable to that reported for other multi-modal digital pathology models, although direct comparisons are limited by differences in cohort composition, endpoint definition, and treatment era(Howard et al., 2023; Witowski et al., 2026).

These findings support the potential clinical validity of CTX as a prognostic tool in postmenopausal, node-positive, HR+ breast cancer. Because CTX derives prognostic information from routinely collected H&E-stained slides without requiring additional tissue sectioning or sequencing, it could provide a scalable and tissue-sparing alternative to genomic assays, which is particularly relevant in resource-limited settings and for patients with limited residual tissue(Howard et al., 2023; Shamai et al., 2026; Witowski et al., 2026).

The chemotherapy-benefit findings are more preliminary. While the treatment-by-biomarker interaction for M-benefit was directionally consistent with predictive ability, it did not reach statistical significance and we combined the two chemotherapy arms rather than restricting the comparison to tamoxifen alone versus sequential CAF-T. Therefore these results should be considered hypothesis-generating rather than confirmatory. This analysis was constrained by modest sample size and a substantial imbalance between the ET (n = 167) and CET (n = 510) arms, which reduces statistical power to detect a treatment-by-biomarker interaction. To preserve power, we combined patients from both chemotherapy arms into a single CET group. This choice departs from the original RS reanalysis (Albain et al., 2010) and a recent multi-modal AI evaluation (Speers et al., 2026), which restricted their predictive analyses to the CAF-T arm. However, both arms contribute treatment-effect information relative to the endocrine-only comparator.

Independent support for the predictive potential of CTX comes from a separate, pre-specified validation of CTX in the TAILORx trial, in which the treatment-by-biomarker interaction for CTX was statistically significant (p = 0.001) among randomly assigned patients with an RS between 11 and 25(Chan et al., 2026). That cohort was substantially larger, with more patients randomized to ET. The direction of the interaction there matches what we observed in this study, which suggests our non-significant result may reflect limited power rather than absence of a true effect. Importantly, TAILORx enrolled node-negative patients, and thus does not provide node-positive-specific validation. Further evaluation in node-positive populations, including contemporary trials such as RxPONDER, will be important. In addition, SWOG-8814 predates taxane-based chemotherapy, aromatase inhibitors, HER2 therapies, CDK4/6 inhibitors, and routine HER2 testing. Prospective validation in contemporary cohorts receiving standard-of-care treatment will therefore be necessary before CTX can be considered for treatment-selection decisions.

This study has several limitations. First, it relied on archival tissue from a trial that accrued participants between 1989 and 1995; prolonged storage may have affected staining quality and tissue morphology. Second, digitized slides were available for only 46% of the original intention-to-treat population, limiting statistical power and introducing the potential for selection bias. Third, the treatment context of SWOG-8814 differs substantially from contemporary practice: CAFT is no longer used in contemporary practice given evidence of inferior efficacy relative to sequential administration (Albain et al., 2009) and the endocrine therapy consisted of tamoxifen rather than an aromatase inhibitor. Fourth, the exploratory analysis of CET benefit was limited by the relatively small number of patients randomized to endocrine therapy alone and was likely underpowered to detect a treatment-by-biomarker interaction. These factors should be considered when interpreting the generalizability and predictive findings of this analysis.

In conclusion, in this blinded external validation of SWOG-8814, Ataraxis Breast CTX provided robust prognostic information for DFS in postmenopausal women with node-positive HR+ breast cancer. CTX also demonstrated a numerically different treatment effect according to predicted benefit from CET, although the treatment-by-biomarker interaction did not reach statistical significance. These findings support CTX as a promising, tissue-sparing prognostic approach and provide a rationale for prospective validation of its predictive performance in contemporary, adequately powered cohorts.

## Funding Information

The SWOG-8814 study and biospecimens from it were funded by the National Cancer Institute (NCI), part of the National Institutes of Health (NIH). The study was led by SWOG Cancer Research Network and was conducted through the NCI’s National Clinical Trials Network (NCTN). Support for this work was provided under NIH/NCI award numbers U10CA180888, U10CA180819, and U24CA196175. Support for biospecimen data processing and analysis was also provided by The Hope Foundation for Cancer Research. This prespecified analysis was funded by Ataraxis AI.

## Statement of Conflicts of Interest

CT, DB JP, KZ, JC, CL, AM, JB, LB, BM, JW, KJG are equity holders of Ataraxis AI. DFH has no direct conflicts for this manuscript. DFH reports research support within the last 12 months provided to his institution, naming him as the principal investigator, from Astra Zeneca, Cepheid, Menarini Silicon Biosystems (MSB), and Pfizer. The University of Michigan (UM) holds patent US 8,790,878 B2 for which D.F.H. is designated as inventor, and which was licensed to MSB with annual royalties paid to UM and DFH, although this agreement expired on 1/1/2023. DFH holds stock options from InBiomotion, CellWorks, and Manta Cares. In the last 24 months, DFH has served as a consultant or on advisory boards for Artera AI Aurexo Therapeutics, Biotheranostics, Cepheid, CellWorks, Delphi Diagnostics, Manta Cares, Milagen, Myriad, Onchilles Pharma, Prism Genomics Medicine, Spotlight Diagnostics, Sysmex, and Veracyte.

## Data Availability

The data is not publicly available due to institutional and ethical constraints. Access to data from the SWOG8814 trial can be requested directly from SWOG.

## Supplemental Tables

**Table S1.** Cox proportional hazards model performance for OS.

| Group | Model | Hazard Ratio | 95% CI | p-value |
| --- | --- | --- | --- | --- |
| **All patients** | H alone | 1.40 | 1.25-1.57 | < 0.001 |
|  | H + clinical features | 1.32 | 1.15-1.51 | < 0.001 |
|  | M alone | 1.66 | 1.47-1.87 | < 0.001 |

H denotes histology score; M, multi-modal score.

**Table S2.** Discriminatory performance for OS.

| Group | Score | Harrell's C-index | 5-Year AUROC |
| --- | --- | --- | --- |
| **All patients** | H-prognostic | 0.60 (0.56-0.64) | 0.63 (0.57-0.69) |
|  | M-prognostic | 0.67 (0.63-0.71) | 0.73 (0.67-0.78) |

H denotes histology score; M, multi-modal score.

## Notes

### Clinical Trial

NCT00929591

### Author Declarations

The SWOG8814 trial was approved by the National Cancer Institute's Central Institutional Review Board and the local institutional review board at each participating site, and all patients provided written informed consent, including consent for the use of tissue for future research. De-identified data were provided by SWOG under a data use agreement.

